# Effects of contrast-medium and vertebral measurement level on computed tomography-based body composition parameters of skeletal muscle and adipose tissue

**DOI:** 10.1101/2024.12.04.24318458

**Authors:** Nobuhiko Akamatsu, Wataru Gonoi, Shouhei Hanaoka, Shohei Inui, Mariko Kurokawa, Satoru Taguchi, Kotaro Sugawara, Haruki Kume, Osamu Abe

## Abstract

**Objective:** To comprehensively evaluate the effects of contrast-medium administration and measurement level (L3 and L1) on computed tomography (CT) derived body composition parameters, promising prognostic factors for various diseases.

**Methods:** 203 dynamic contrast-enhanced CT examinations, including unenhanced (phase 0) and early arterial, late arterial, portal, and equilibrium phases (phases 1-4, respectively), were retrospectively enrolled in the study. Areas and mean densities of skeletal muscle (SMA and MSMD), subcutaneous adipose tissue (SATA and MSATD), and visceral adipose tissue (VATA and MVATD) were measured at L3 and L1 levels across five phases (phase 0–4). Measurement changes among phases and levels were assessed statistically.

**Results:** The rates of SMA, SATA, and VATA on phase 1/2/3/4 compared to those on phase 0 were as follows: L3 SMA +1.1%/+2.1%/+2.8%/+3.5%; L1 SMA, +2.0%/+2.7%/+3.0%/+3.7%; L3 SATA −0.4%/−1.9%/−2.9%/−4.7%; L1 SATA −0.3%/−2.2%/−3.5%/−6.1%; L3 VATA −7.5%/−17.8%/−20.0%/−22.2%; L1 VATA −8.5%/−20.1%/−22.6%/−23.7%. Differences of MSMD, MSATD, and MVATD on phase 1/2/3/4 between those on phase 0 were as follows (Hounsfield Units): L3 MSMD, +2.1/+5.8/+7.8/+9.6; L1 MSMD, +2.9/+7.7/+9.1/+9.8; L3 MSATD, +1.2/+3.7/+4.8/+6.2; L1 MSATD, +1.2/+3.8/+4.7/+5.6; L3 MVATD, +1.5/+4.1/+5.2/+6.1; L1 MVATD, +1.9/+4.9/+5.8/+6.3. The values between L3 and L1 showed a linear solid correlation (coefficient of determination, 0.950–0.999), suggesting interchangeability.

**Conclusion:** Changes in body composition parameters measured in various contrast phases and two dominant body levels were comprehensively elucidated, promoting the interchangeability of cohorts with mixed CT conditions.

## Introduction

Previous studies have revealed that sarcopenia, characterized by the quality and quantity of the skeletal muscle, subcutaneous adipose tissue, and visceral adipose tissue composing the human body, is associated with treatment outcomes and the prognosis of various diseases.

A decrease in skeletal muscle mass has been associated with a higher incidence of falls and fractures, as well as poor outcomes in various conditions, including pneumonia, cardiovascular catheterization, and pancreatitis [1–5]. It is also linked to poor prognosis of various cancers in the head and neck, lung, gastrointestinal tract, liver, bile duct, pancreas, and urinary tract [1–8]. For quantitative measurement of skeletal muscle mass, computed tomography (CT) is frequently used as a simple and minimally invasive method [9]. In particular, the skeletal muscle area (SMA) measured at the L3 vertebral level highly correlates with total body skeletal muscle mass. Shen et al. reported a correlation coefficient of 0.924 between SMA at the L3 level and total body skeletal muscle mass [10]. An L3-level skeletal muscle index (SMI, calculated as SMA divided by the square of height) is the most predominant indicator of total body skeletal muscle mass that considers personal physique [8].

In contrast, myosteatosis, in which adipose tissue accumulates within skeletal muscle, reducing muscle quality, has been reportedly associated with all-cause mortality in health check-up participants and the prognosis of multiple cancers [11,12]. Skeletal muscle density (as measured by CT value) has been reported to reflect the abundance of adipose tissue within the muscle [11].

Increased visceral adipose tissue mass is associated with hypertension, dyslipidemia, insulin resistance, and hyperglycemia, while increased subcutaneous adipose tissue is associated with metabolic diseases [13,14]. Furthermore, associations between visceral and subcutaneous adipose tissue amounts and the prognosis of multiple cancers have been reported [13–15]. It is reported that the adipose tissue area at a transaxial image at a lumbar vertebral level strongly correlates with total body adipose tissue mass [10]. The vertebral level visceral and subcutaneous adipose tissue index (areas divided by the square of height to compensate for physical difference) are the major representative indicators of total body visceral and subcutaneous adipose tissue mass [10,15,16].

Evidence has accumulated for the usefulness of measuring body composition parameters, including areas or densities of skeletal muscle and subcutaneous or visceral adipose tissue, with various clinically meaningful cutoff criteria proposed for these parameters. However, the methodology for its measurement still requires further elaboration and refinement. These tissues are usually contoured on CT images using established specific CT value thresholds. The problem is that most previous studies have not considered the effect of contrast medium administration and post-contrast timing on measured CT values and calculated parameters.

As the CT values of muscle and adipose tissues increase after contrast agent administration and by post-contrast phases [17], the measurement would also change when using the established specific CT value thresholds. Some previous studies, with limited sample sizes, contrast conditions, and measured parameters, reported minimal changes in SMA and subcutaneous adipose tissue (SATA) due to contrast agent administration. In contrast, changes in visceral adipose tissue area (VATA) were relatively significant [18–23].

Another limitation is that the L3 vertebral level is only sometimes included in upper abdominal CT scans and is absent in chest CT scans. Conversely, the L1 vertebral level is consistently within the imaging range of both modalities. While the L3 level is the standard for body composition analysis, the availability of L1-derived metrics could be valuable for patients when L3 imaging is unavailable. Previous studies have also suggested the usefulness of L1-based body composition parameters [24,25].

To address the above issues and promote data interchangeability, this study aimed to comprehensively evaluate the effects of multi-phase contrast-medium administration and the measurement level (L3 and L1) on CT-derived body composition parameters, anticipating its potential as a promising prognostic factor for various diseases.

## Materials and Methods

### Ethics

This study was approved by the Ethics Committee/Clinical Research Review Board of our university hospital (Approve Number, 2561). As this was a retrospective study without intervention, it was determined that obtaining patient consent and registration in a clinical research registry were not necessary.

### Study Population

This retrospective study first enrolled 338 consecutive upper-abdominal dynamic contrast-enhanced CT examinations using a five-phase contrast-enhance protocol designated for hepatobiliary diseases at our university hospital between April 4, 2022, and May 19, 2022. We excluded duplicate examinations of the same patient (n=16), examinations lacking the level of the first and the third lumbar vertebra (L3 and L1 levels) or any of the five phases (n=74), examinations with skeletal muscle deformities or metal implants or other medical devices due to operations (n=8), and examinations with severe edema or emaciation where skeletal muscles were difficult to contour (n=37). The remaining 203 examinations were finally included in the analysis.

### CT Protocol

All CT scans were performed using the following CT scanners: Aquilion ONE (Canon Medical Systems, Tochigi, Japan), Aquilion PRIME (Canon Medical Systems), and Discovery 750 (GE Healthcare, Chicago, US). Scanning parameters were as follows for all scanners: tube voltage, 120 kVp; tube current, determined using auto exposure control. For image reconstruction, filtered back projection was used for Aquilion ONE and Aquilion PRIME, while Adaptive Statistical Iterative Reconstruction was employed for Discovery 750. The imaging phases were as follows: non-contrast (phase 0), early arterial phase (phase 1, pre-scan breath-hold instructions were provided when the abdominal aorta at the level of the diaphragm reached 200 HU), late arterial phase (phase 2, 15 seconds after the phase 1), portal venous phase (phase 3, 70 seconds after the start of contrast administration), and equilibrium phase (phase 4, 180 seconds after the start of contrast administration), totaling five phases. Transaxial images with a slice thickness of 5 mm were used for analysis.

### Image Analysis

We used a deep learning-based custom software implemented on the CIRCUS version 1.6.0 platform (http://circus-project.net), a computer-aided diagnosis tool as utilized in the previous studies [7,8,26]. For each CT examination, including five imaging phases, two transaxial planes at the middle of the first and third vertebrae (L3 and L1 level), totaling ten images per examination, were selected automatically using the software. Then, the software automatically segmented all the selected images into skeletal muscle, subcutaneous adipose tissue, visceral adipose tissue, and others, using the thresholds presumed to have been proposed for non-contrast CT as follows, according to previous studies: skeletal muscle, −29 to 150; subcutaneous adipose tissue, −190 to −30 H.U.; visceral adipose tissue, −150 to −50 HU (Fig. 1) [15,16,27,28]. The subcutaneous adipose tissue area was defined between the skin and the outer contour of the skeletal muscles and met the threshold mentioned above. The visceral adipose tissue area was designated as the inner side of the skeletal muscles and met the threshold. Board-certified radiologists (W.G., N.A, S.I.) closely reviewed all the selected slices and segmented images and manually corrected the software’s output until they were correct. Using the segmented images, the following six parameters of body composition were extracted for each image: SMA (cm^2^); SATA (cm^2^); VATA (cm^2^); mean skeletal muscle density (MSMD) (HU); mean subcutaneous adipose tissue density (MSATD) (HU); mean visceral adipose tissue density (MVATD) (HU).

**Fig. 1.**
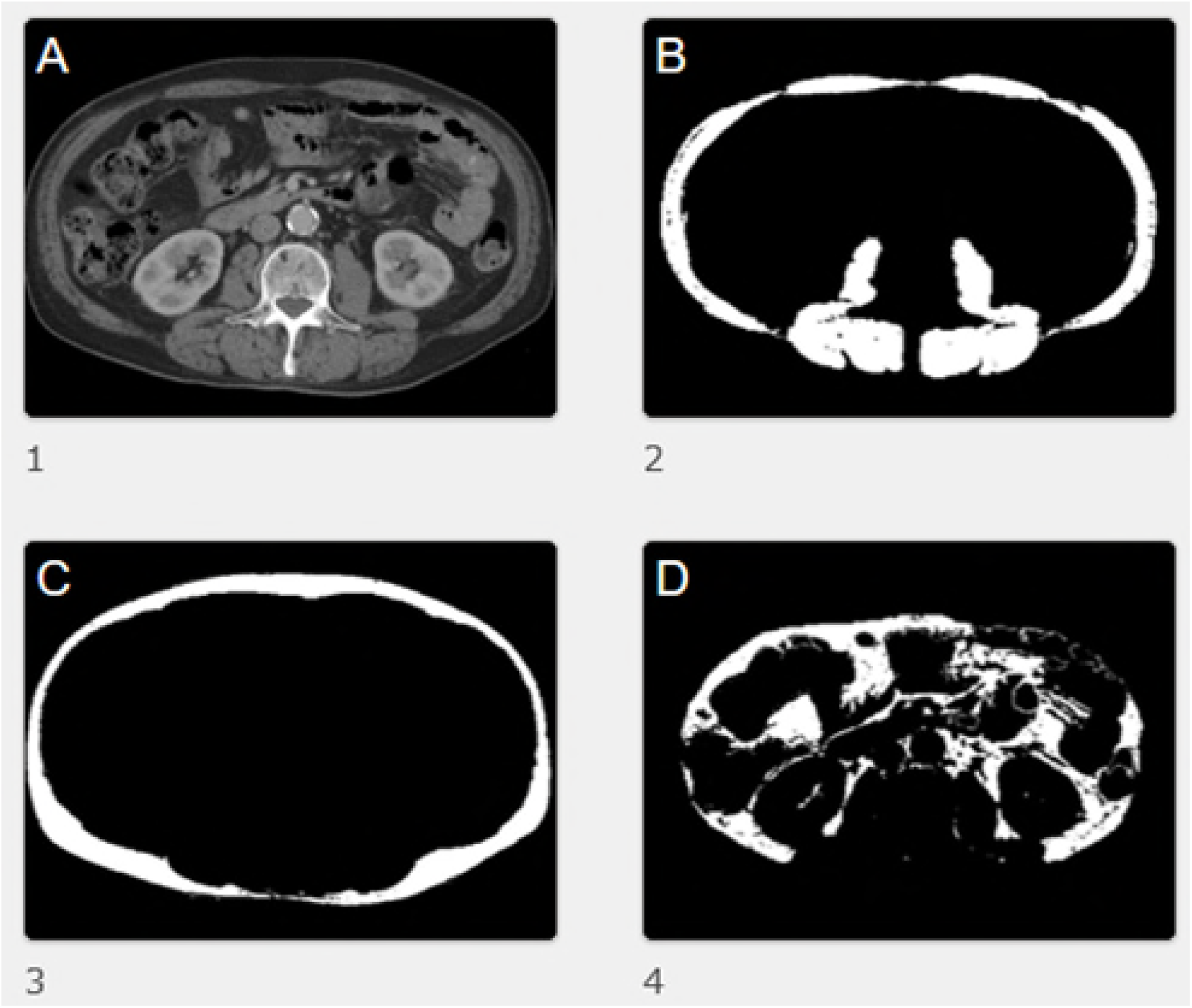
An example of body parameter segmentation analysis results from the deep-learning program: (A) Original portal phase enhanced CT image at the L3 level before processing; (B) Results of segmentation of the skeletal muscle; (C) the subcutaneous adipose tissue;(D) and the visceral adipose tissue.

### Statistical Analysis

For each indicator (SMA, SATA, VATA, MSMD, MSATD, and MVATD) at the L3 and L1 levels, changes from phase 0 to phases 1/2/3/4 were calculated, respectively. The trend of these changes over imaging phases was tested using the Wilcoxon signed-rank test. Scatter plots with regression lines and coefficient of determination and the Spearman’s correlation coefficients were employed to test the correlation of the indicators between the L3 and L1 levels. Statistical analyses were performed using EZR ver.1.61 (Jichi Medical University Saitama Medical Center, Tochigi, Japan), a modified statistical software derived from R commander (The R Foundation for Statistical Computing, Vienna, Austria).

## Results

The backgrounds of the 203 cases included in the analysis were as follows: age, median 69 (interquartile range, 57-78); male/female, 132/71; and body weight, median 62 (interquartile range, 53-68).

Fig. 2 depicts the change ratios of contrast phases 1/2/3/4 (early arterial/late arterial/portal/equilibrium phases) to phase 0 (unenhanced) for SMA, SATA, and VATA at L3 and L1 levels. The rates of SMA, SATA, and VATA on phase 1/2/3/4 compared to those on phase 0 were as follows: L3 SMA, +1.1%/+2.1%/+2.8%/+3.5%; L1 SMA, +2.0%/+2.7%/+3.0%/+3.7%; L3 SATA −0.4%/−1.9%/−2.9%/−4.7%; L1 SATA −0.3%/−2.2%/−3.5%/−6.1%; L3 VATA −7.5%/−17.8%/−20.0%/−22.2%; L1 VATA −8.5%/−20.1%/−22.6%/−23.7%. In summary, the SMA showed a monotonic and slight increase over the phases, while SATA and VATA showed a slight decrease with significance over the phase at both L3 and L1 vertebral levels. Wilcoxon signed-rank test elucidated significant differences in SMA, SATA, and VATA among phases 0/1/2/3/4 at each level, respectively. Wilcoxon signed-rank test revealed significant differences between most couples of imaging phases for SMA, SATA, and VATA at both L3 and L1 levels.

**Fig. 2.**
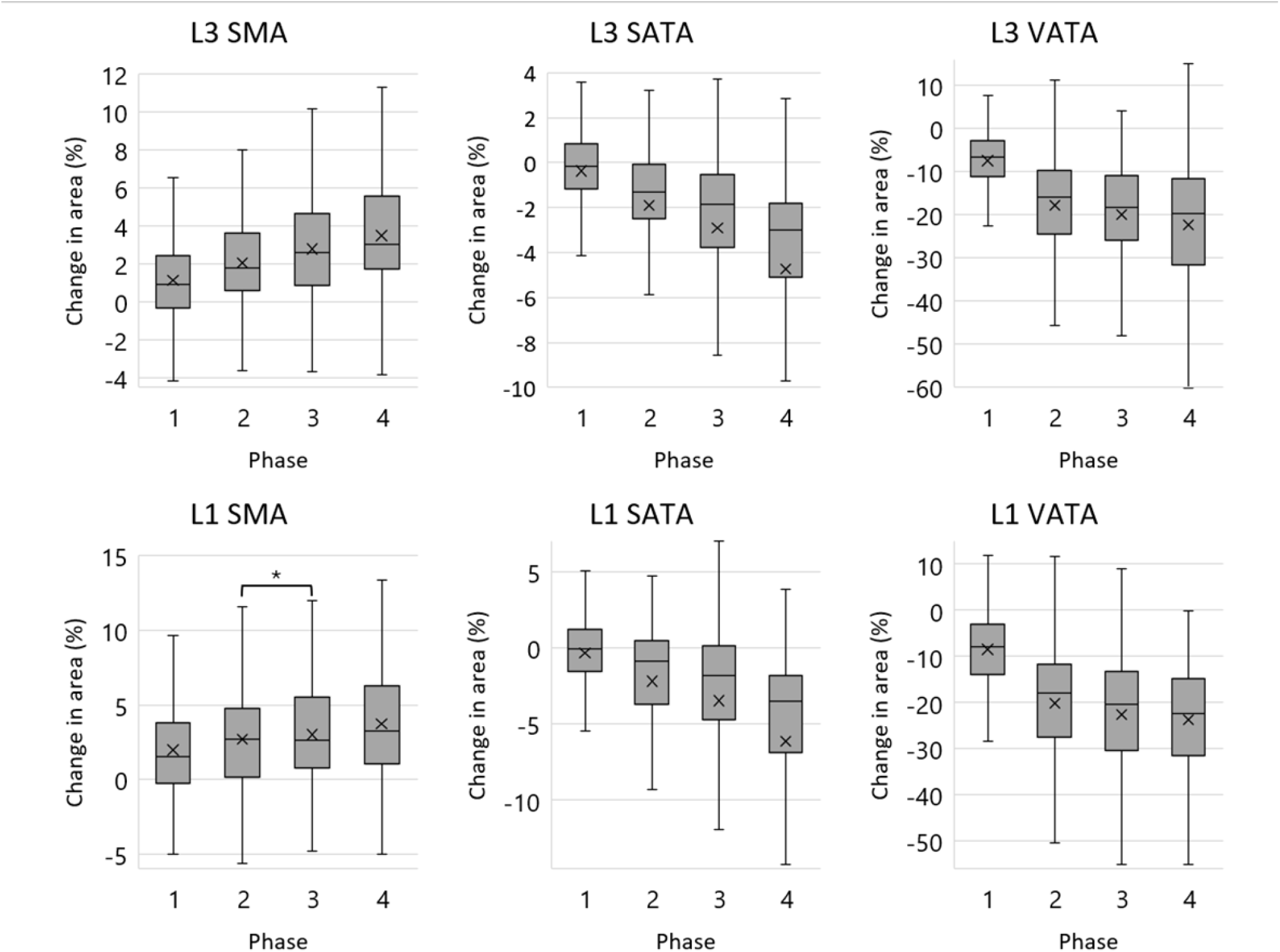
The box plots display the change ratios (%) of early arterial/late arterial/portal/equilibrium phases (phases 1/2/3/4) to unenhanced phase (phase 0) for skeletal muscle area (SMA), subcutaneous adipose tissue area (SATA), and visceral adipose tissue area (VATA) at the L3 and L1 levels are depicted. ‘x’ represents the mean value. ‘*’ represents statistically insignificant by the Wilcoxon signed-rank test (*p* = 0.063), and the other pairs were all statistically significant (*p* < 0.001 for the other pairs).

Fig. 3 shows the CT value changes in MSMD, MSATD, and MVATD at the L3 and L1 levels for phases 1/2/3/4 compared to phase 0. Differences of MSMD, MSATD and MVATD on phase 1/2/3/4 between those on phase 0 were as follows (HU): L3 MSMD, +2.1/+5.8/+7.8/+9.6; L1 MSMD, +2.9/+7.7/+9.1/+9.8; L3 MSATD, +1.2/+3.7/+4.8/+6.2; L1 MSATD, +1.2/+3.8/+4.7/+5.6; L3 MVATD, +1.5/+4.1/+5.2/+6.1; L1 MVATD, +1.9/+4.9/+5.8/+6.3. In summary, the MSMD, MSATD, and MVATD in contrast-enhanced CT showed a monotonic increase over time (over phase) at both L3 and L1 vertebral levels. Wilcoxon signed-rank test revealed significant differences between all ten couples of five imaging phases for L3 MSMD, L1 MSMD, L3 MSATD, L1 MSATD, L3 MVATD, and L3 MVATD, respectively (*p* < 0.001 for all).

**Fig. 3.**
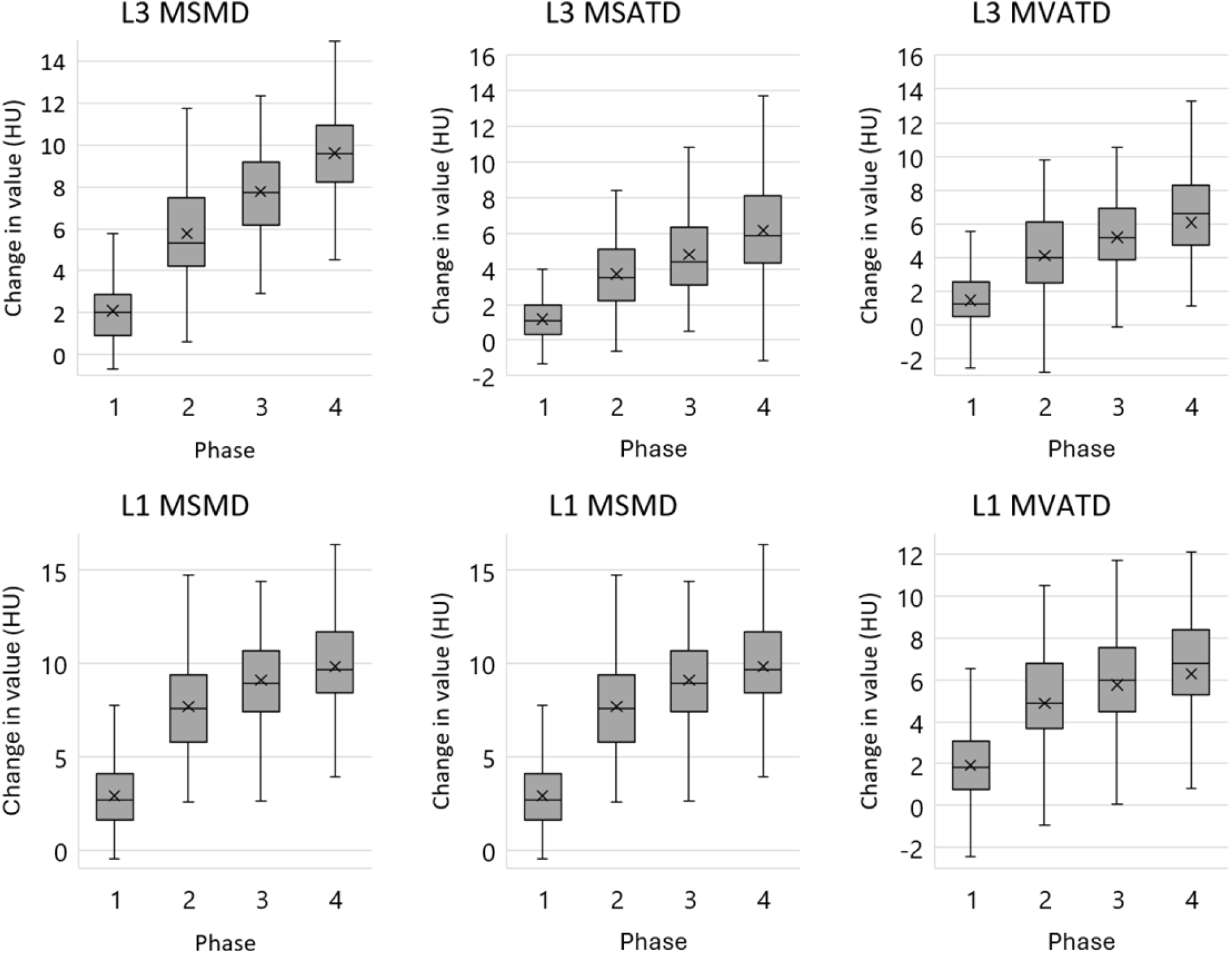
The box plots display the CT value changes (HU) in mean skeletal muscle density (MSMD), mean subcutaneous adipose tissue density (MSATD), and mean visceral adipose tissue density (MVATD) at the L3 and L1 levels for early arterial/late arterial/portal/equilibrium phases (phases 1/2/3/4) to unenhanced phase (phase 0). ‘x’ represents the mean value. The Wilcoxon signed-rank test revealed all the pairs of imaging phases in each parameter were statistically significant (p < 0.001 for all).

Scatter plots calculated to test a correlation between L3 and L1 levels for SMA, SATA, VATA, MSMD, MSATD, and MVATD in each imaging phase showed a linear solid correlation with a high coefficient of determination ranging from 0.950 to 0.999 in all imaging phases (Fig. 4 and Fig. 5). Spearman’s rank correlation coefficients ranged from 0.925 to 0.948 with p-values of <0.001 for all the pairs of L3 and L1 values in each body composition parameter. The measurement values between L3 and L1 can be converted interchangeably using the equations of the linear regression lines passing through the origin, as shown in Fig. 4 and Fig. 5.

**Fig. 4.**
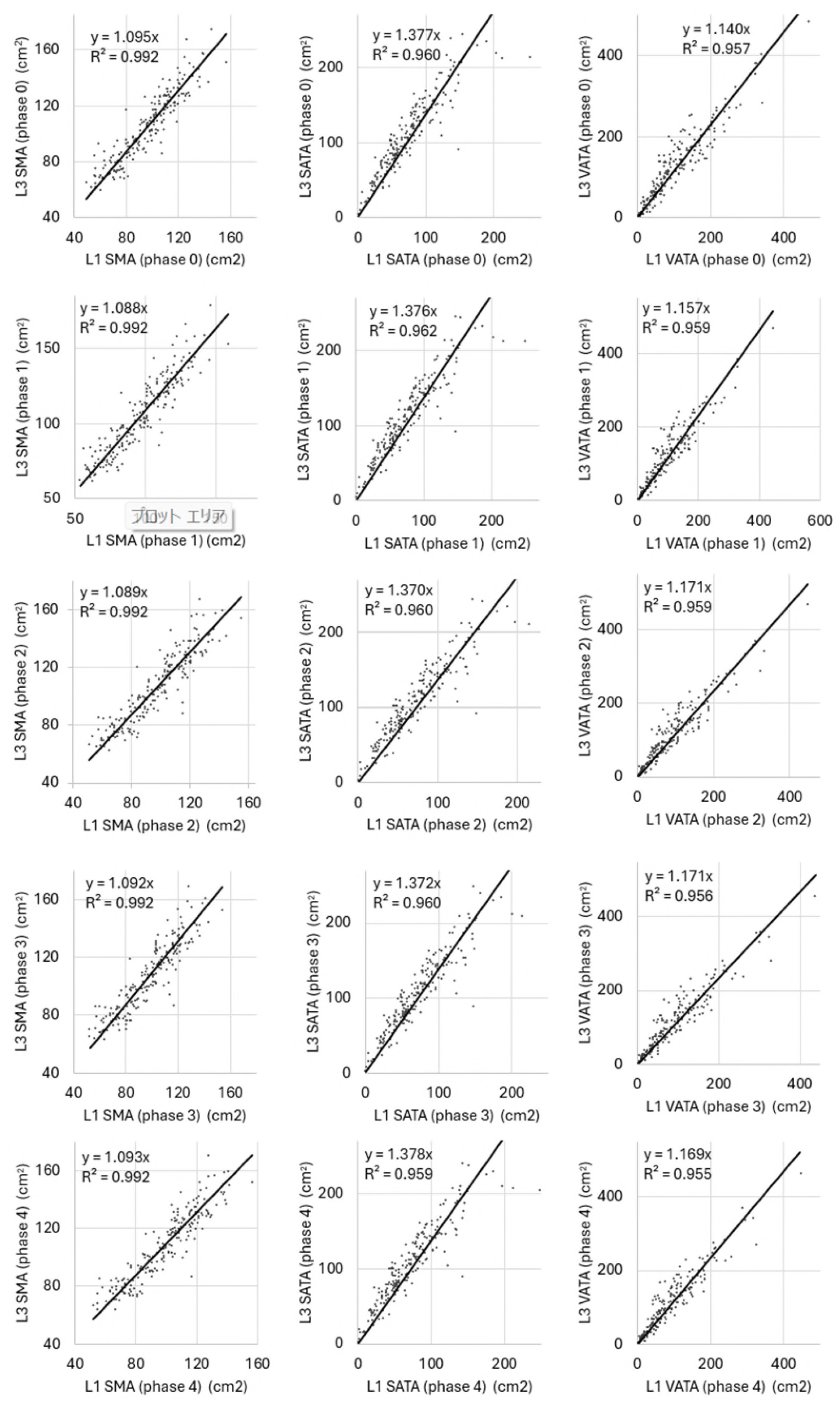
The scatter plots display a correlation between the L3 and L1 levels for skeletal muscle area (SMA), subcutaneous adipose tissue area (SATA), and visceral adipose tissue area (VATA) in unenhanced/early arterial/late arterial/portal/equilibrium phases (phases 0/1/2/3/4) depicted along with their regression equation and coefficient of determination. The SMA, SATA, and VATA values between the L3 and L1 levels strongly correlated linearly in all imaging phases.

**Fig. 5.**
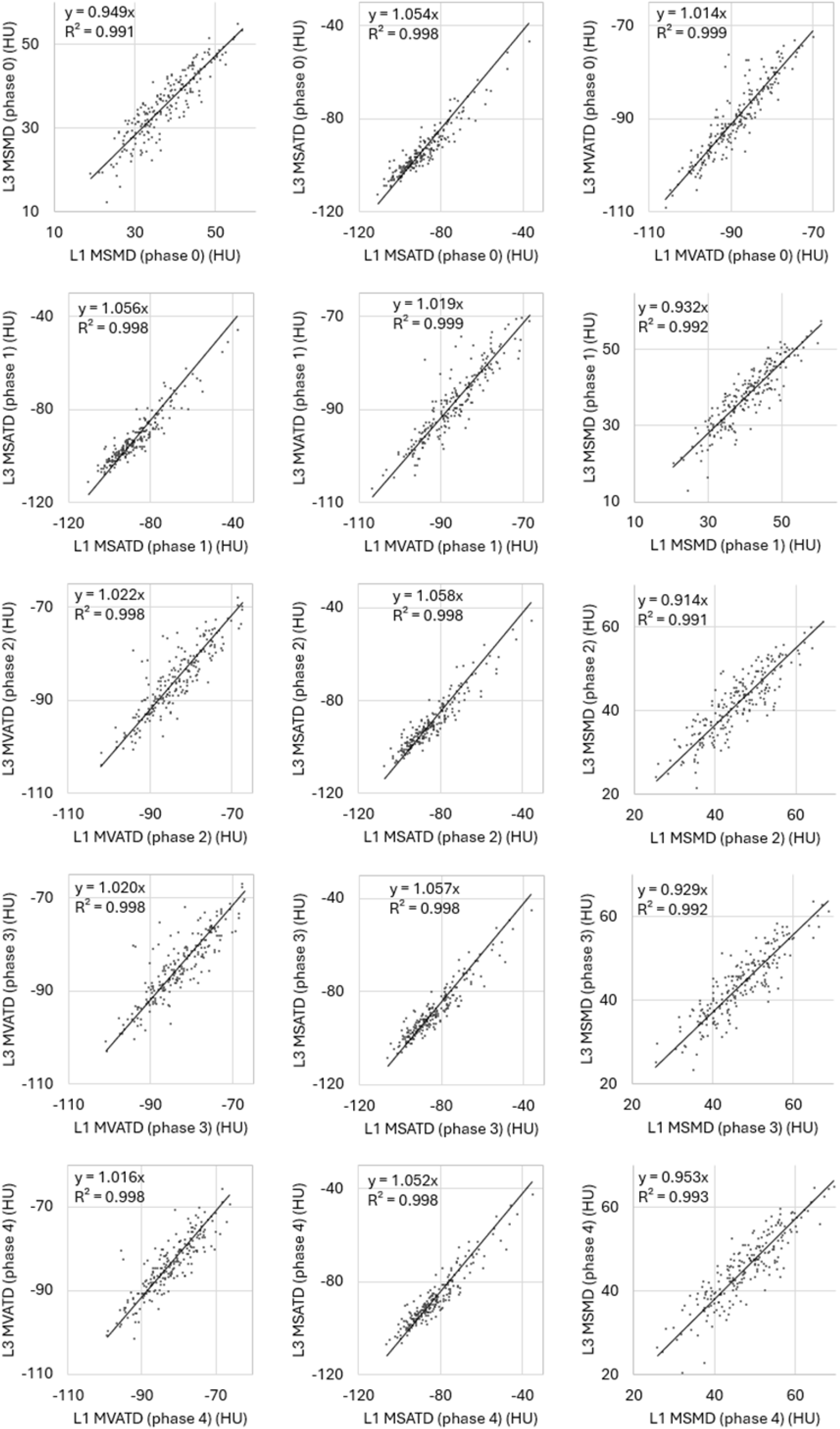
The scatter plots display a correlation between L3 and L1 levels for mean skeletal muscle density (MSMD), mean subcutaneous adipose tissue density (MSATD), and mean visceral adipose tissue density (MVATD) in unenhanced/early arterial/late arterial/portal/equilibrium phases (phases 0/1/2/3/4) depicted along with their regression equation and coefficient of determination. The MSMD, MSATD, and MVATD values between the L3 and L1 strongly correlated linearly in all imaging phases.

## Discussion

In the present study, we comprehensively measured several body composition parameters on five-phase dynamic-enhanced CT at L3 and L1 levels, revealing the detailed effects of contrast medium administration and a strong correlation between L3 and L1 measurements. Furthermore, utilizing the aforementioned area change ratio and CT value change might enable accurate estimation of measurement values at different contrast phases or vertebral levels through numerical transformation.

SMA increased monotonously following contrast agent administration over time at L3 and L1 vertebral levels, while SATA and VATA decreased monotonously. MSMD, MSATD, and MVATD showed monotonic increases at L3 and L1 levels after contrast agent administration. These results reflect the intravenously administered contrast agent gradually expanding its distribution from the intravascular space to the extravascular space and interstitial compartments over time [29]. While changes in SMA and SATA were minimal, VATA showed a significant decrease following contrast agent administration. This could be because visceral adipose tissue is adjacent in a wide range to parenchymal organs that exhibit more significant contrast enhancement compared to subcutaneous adipose tissue, and therefore, VATA was underestimated due to the partial volume effect.

Regarding skeletal muscle parameters, a previous study with 38 patients reported a minimal increase in SMA (+1.9%) and MSMD (+1.4 HU) in the arterial phase compared to the non-contrast phase at the L3 level [23]. Another study with 89 patients reported monotonous increases in SMA (arterial/portal/delayed phases, +0.5%/+1.5%/+1.8%) and MSMD (+6.2 HU/+11.5 HU/+14.2 HU) at the L3 level after dynamic contrast-medium administration [27]. Another study with 316 healthy patients also reported increases in SMA (early/late arterial phases, +0.8%/+1.7%) and MSMD (+5.5 HU/+8.0 HU) at the L3 level [30]. All these studies showed slight increases in SMA and MSMD over time following contrast agent administration, which was consistent with our findings.

Regarding subcutaneous and visceral adipose tissues, a previous study with 31 patients reported abdominal to pelvic subcutaneous and visceral adipose tissue volumes decreased by 7.3% and 7.7% after contrast medium administration, respectively [20]. Another study reported a 9.4% decrease in SATA and a 25.4% decrease in VATA at the L1 level after contrast enhancement with an atypical CT protocol for kidney donors [31]. Those results for adipose tissues were consistent with our findings.

As for inter-level comparison, in the present study, SMA, SATA, and VATA showed strong correlations between the L3 and L1 levels across all imaging phases. This suggests that SMA, SATA, and VATA at the L1 level, similar to those at the L3 level, could potentially be used for prognostic prediction in cancer patients. Regarding previous studies on the correlation of SMA at different levels, a non-contrast MRI study with 155 cirrhosis patients demonstrated a strong correlation in SMA between the L3 and L1 levels, proposing an equation for SMA between the levels [24]. Another non-contrast CT study with 131 patients showed solid correlations between the L3 and L1 levels for SMA and MSMD [25]. Another large-scale study with 1677 patients measured VATA and MVATD at multiple thresholds for the T10 to L4 levels [19].

The strength of the present study over previous investigations is its comprehensive analysis of clinically relevant body composition parameters across multiple contrast-enhanced imaging phases at the L3 and L1 levels, which are known for their prognostic value. With a larger sample size than previous studies, our findings on changes in area and density values could help integrate body composition metrics from different imaging protocols in heterogeneous populations, improving prognostic predictions.

The present study has a few limitations. The prevalence of liver disease in the present cohort could have been high, and the ethnicity of the participants was Asian.

In conclusion, the measures of body composition parameters vary among various phases of contrast-medium administration and measurement levels. The conversion factors provided by the present results enable more concise body parameter analyses among cohorts with mixed imaging protocols.

## Data Availability

All data produced in the present study are available upon reasonable request to the authors

## Acknowledgment

Nobuhiko Akamatsu and Shohei Inui contributed equally to the manuscript.

All authors have seen and approved the manuscript.

